# All-cause mortality and causes of death in the Swiss Hepatitis C Cohort Study

**DOI:** 10.1101/2019.12.13.19014837

**Authors:** Maroussia Roelens, Barbara Bertisch, Darius Moradpour, Andreas Cerny, Nasser Semmo, Patrick Schmid, Beat Müllhaupt, Olivier Clerc, David Semela, Christoph Junker, Francesco Negro, Olivia Keiser, for the Swiss Hepatitis C Cohort Study

## Abstract

**Background & Aims:** With the introduction of direct-acting antiviral agents (DAA), mortality rates and causes of death among persons with hepatitis C virus (HCV) infection are likely to change over time. However, the emergence of such trends may be delayed by the relatively slow progression of chronic hepatitis C. To date, detailed analyses of cause-specific mortality among HCV-infected persons over time remain limited.

**Methods:** We evaluated changes in causes of death among the Swiss Hepatitis C Cohort Study (SCCS) participants, from 2008 to 2016. We analysed risk factors for all-cause and cause-specific mortality, accounting for changes in treatment, fibrosis stage and use of injectable drugs over time. Mortality ascertainment was completed by linking lost-to-follow-up participants to the Swiss Federal Statistical Office (SFSO) death registry.

**Results:** We included 4,700 SCCS participants, of whom 478 died between 2008 and 2016. Linkage to the SFSO death registry substantially improved the information on causes of death (from 42% of deaths with unknown cause to 10% after linkage). Leading causes of death were liver failure (crude death rate 4.4/1000 person-years), liver cancer (3.4/1000 p-yrs) and non-liver cancer (2.8/1000 p-yrs), with an increasing proportion of cancer-related deaths over time. Cause-specific analysis showed that persons with sustained virologic response (SVR) were less at risk for liver-related mortality than those never treated or treated unsuccessfully.

**Conclusions:** Although the expected decrease in mortality is not yet observable, causes of death among HCV-infected persons evolved over time. With the progressive widening of guidelines for DAA use, liver-related mortality is expected to decline in the future. Continued monitoring of cause-specific mortality will remain important to assess the long-term effect of DAA and to design effective interventions.

**Lay summary:** Leading causes of death among persons with hepatitis C virus (HCV) infection in the Swiss Hepatitis C Cohort study evolved over the past years, with an increasing proportion of cancer-related deaths. The positive impact of new potent anti-HCV drugs on mortality among HCV-infected persons is not yet observable, due to both the slow progression of chronic hepatitis C and the progressive relaxation of guidelines for the use of those new drugs.

## Background

Hepatitis C virus (HCV) infection is a leading cause of liver-related mortality [1, 2], responsible for about 399,000 deaths worldwide in 2016. With the introduction of potent direct-acting antiviral agents (DAA), the risk of morbidity and mortality for HCV infected persons has decreased substantially [3]. It is likely that causes of death are changing over time and with age, since HCV infected people may become increasingly at risk of dying from non-liver-related causes, including non-liver-related malignancies or cardiovascular diseases [4]. However, due to the slow progression of chronic hepatitis C, such trends may be slow to emerge. Some studies predict a rise in liver-related deaths to continue for another decade [5, 6], meanwhile the number of HCV-related deaths are stable or increasing in many settings [7].

Various studies have analysed mortality among HCV-infected persons. In the United States (US), several studies have shown that HCV-related deaths disproportionally affect persons born between 1945 and 1965 [8, 9]. Both a comparison between the Chronic Hepatitis Cohort Study and the official death certificates in the US [9], and a comparison between the Swiss Hepatitis C Cohort Study (SCCS) and the death certificates from the Swiss Federal Statistical Office (SFSO) [7] showed that under-reporting of HCV infection on death certificates is quite common. In both the US and Switzerland, HCV-related mortality increased significantly between the late 1990s and the early 2000s, whereas human immunodeficiency virus (HIV)-related mortality decreased, and hepatitis B virus (HBV)-related mortality remained relatively stable [7, 8]. In New South Wales, Australia, HCV notifications were linked to the death registry [1, 2]. When comparing mortality between HCV-infected persons and the general population, both liver-related and drug-related death rates were about 15-16 times higher in Australians with HCV. In HCV mono-infected persons, liver-related deaths increased considerably with older age, and drug-related death rates were higher in younger age groups [1, 2]. In this population, liver-related mortality remained stable between 1997 and 2006, and drug-related mortality was relatively constant between 2002 and 2006 [1, 2]. More recently, a study based on the US Veterans Affairs Hepatitis C Clinical Case Registry showed that, among HCV-infected patients treated with DAA, successful treatment substantially reduced mortality, both in the presence and absence of advanced liver disease [10, 11].

Nevertheless, few studies have analysed detailed patterns of cause-specific mortality among HCV-infected persons. We therefore investigated the risk factors and time trends for all-cause and cause-specific mortality among HCV-infected persons in the SCCS. We improved mortality ascertainment by linking patients lost-to-follow-up to death certificates from the SFSO.

## Methods

### The Swiss Hepatitis C Cohort Study (SCCS)

The SCCS is a prospective observational cohort study, established in 2000, that continuously enrols adult patients aged ≥ 18 years in Switzerland. Only persons who are confirmed anti-HCV antibody-positive are included [12]. Eight centres are involved, including all five university hospitals in Switzerland (Basel, Bern, Geneva, Lausanne, Zurich), three large non-university hospitals (Lugano, Neuchâtel and St. Gallen) and some affiliated centres. For all persons in the SCCS, various demographic, psychosocial, clinical, laboratory and treatment data were collected via standardized questionnaires. The questionnaires were completed by physicians or study nurses, during enrolment and annual follow-up visits. The study was approved by all local ethics committees, and all persons provided written informed consent. For deceased persons, the cause of death was coded according to the International Statistical Classification of Diseases and Related Health Problems (ICD-10) [13]. Supplementary information was also provided to specify if the death was related to HCV infection or due to an accident, a suicide or overdose/accidental poisoning.

### Eligibility criteria and definitions

We included all persons enrolled before December 31^st^ 2016 (until when data on causes of death were compiled by the SFSO). We excluded those who died before January 1^st^ 2008, because the new 13-digit social security number (SSN), which was needed for the linkage, became available only in 2008. Patients were lost-to-follow-up (LTFU) if they were not seen since August 2015 and not known to have moved abroad or to have died. Sustained virologic response (SVR) was defined as an undetectable HCV RNA ≥ 12 weeks after the end of antiviral treatment.

### Linkage with the SFSO death registry

The SFSO has monitored causes of death in Switzerland since 1876. Causes of death are coded by the ICD-10 code based on initial, consecutive and concomitant diseases listed on the SFSO death certificate. To complete the SCCS information on mortality, deceased and LTFU persons in the SCCS were linked to the SFSO death registry, by exact record linkage. The linkage was done for all persons who were declared LTFU or dead in the SCCS between January 1^st^ 2008 and December 31^st^ 2016. We used the 13-digit SSN and the date of birth for the linkage when the SSN was available. If the SSN was not available, the linkage was based on the dates of birth and death.

### Coding and comparing causes of death

We classified the main cause of death into mutually exclusive categories (liver cancer, liver failure, non-liver cancer, cardiovascular, unnatural causes, other, unspecified) as described in Table 1. The “other” category includes all deaths with a frequency below 30 persons. Unnatural causes of death include suicide, overdose/accidental poisoning and accident.

**Table 1:** Categories of causes of death.

| Death cause group | Eligibility criteria |
| --- | --- |
| Liver failure (other than liver cancer) | Selected ICD-10 codes:<br>B15 - B19 (viral hepatitis)<br>B94.2 (sequelae of viral hepatitis)<br>K70 (alcoholic liver disease)<br>K71 - K77 (toxic/chronic/other liver disease)<br>T86.4 (liver transplant failure and rejection)<br>Z94.4 (liver transplant status)<br>OR<br>Death identified as physically related to HCV infection according to information obtained at or after a SCCS visit |
| Liver cancer | Selected ICD-10 code: C22 |
| Non-liver cancer | Selected ICD-10 codes:<br>C00 - C21, C23 - C97 (all malignant neoplasms except liver cancer, including leukaemia)<br>D00 - D48 (all in-situ, benign and unknown neoplasms) |
| Unnatural cause | Selected ICD-10 codes:<br>S00 - T88 except T86.4 (injury, poisoning and certain other consequences of external cause)<br>V01 - Y98 (external causes of morbidity and mortality)<br>Z92 - Z99 except Z94.4 (health-service-related factor)<br>OR<br>Death identified as due to accident, suicide or overdose of narcotics according to the SCCS |
| Cardiovascular | Selected ICD-10 codes:<br>I00 - I99 (diseases of the circulatory system) |
| Other | All the deaths with selected ICD-10 code not entering any of the criteria described above |
| Unspecified | No selected ICD-10 code and no complementary information from the SCCS |

For each deceased person in the SCCS, we retained one source of information to classify the cause according to the following rules. For persons linked to the death registry, we preferentially selected ICD-10 codes from the SFSO. When this information was not available, or when the SFSO provided less specific information than the SCCS, we used the ICD-10 code from the SCCS. In case no ICD-10 code was available from the SFSO or the SCCS, we used the supplementary information from the SCCS on causes of death mentioned earlier, specifying if the death was physically related to HCV infection, or due to an accident, a suicide or an overdose/accidental poisoning. All the deceased persons who could not be classified following this procedure were defined “unspecified”. Finally, the cause of death classification was reviewed by an expert clinician who corrected misclassifications of complex cases specific to HCV infection.

For the deceased persons linked to the SFSO death registry, we compared the main causes of death in the SCCS and the SFSO. We measured the interrater agreement via the Kappa statistics in two ways [14]: first by directly comparing the ICD-10 codes for the main cause of death, and then by using the grouped causes of death using the groups in Table 1.

### Statistical analyses

We calculated overall crude mortality rates and for different time periods, for both each cause of death group and all-cause mortality. We also conducted survival analysis, for which follow-up time was calculated from enrolment into the cohort or the starting date of the period of interest (baseline), to the censoring date (which was either the date of death or the last known date of being alive). When the dates of death in the SCCS and SFSO were different, we used the date of the SFSO.

We calculated crude cumulative incidences for each cause of death group, accounting for competing risk (where each cause of death was a competing risk for the other causes), using the R *mstate* package [15]. We then used Cox Proportional Hazard regression, including time-dependent covariables [14, 15], to determine risk factors of mortality. We first analysed all-cause mortality, and then cause-specific mortality, for each of the causes of death (except for “other” and “unspecified”). Analyses were adjusted for sex, baseline age (as a continuous variable with restricted cubic splines [18]), fibrosis score (F1 to F4), history of injection drug use (IDU) (ever, never), and treatment status (never treated, treated with SVR, treated without SVR). For all-cause mortality, we also accounted for the treatment history (never treated, ever treated and received DAA, ever treated but never received DAA). Due to the limited number of deceased patients who received DAA, we could not add this co-variable in the cause-specific analysis. To determine the fibrosis score, we combined the information from liver biopsies, FibroScan^™^ analyses, and any reported cirrhosis during follow-up. Liver stiffness assessed by FibroScan^™^ was converted to Metavir scores (F1: <7.5 kPa, F2: ≥ 7.5 to < 9.5 kPa, F3: ≥ 9.5 to < 12.5 kPa, and F4: ≥ 12.5 kPa) and reported cirrhosis was translated into a Metavir fibrosis score of 4. We made no assumption on the possible evolution of fibrosis over time, and carried the values forward until the next measurement or until the end of the study. We used multiple imputation by chained equations (R *mice* package [19]) to impute missing values of time-dependent covariables (see details in Appendix 1).

For sensitivity analysis, we recalculated the Cox Proportional Hazard models without multiple imputation, excluding all the persons for whom information on treatment type and outcome, fibrosis score, and history of IDU was incomplete at any time over the period of interest.

## Results

### Linkage SCCS – SFSO death registry

A total of 4,700 SCCS participants were followed between January 1^st^ 2008 and December 31^st^ 2016, with 471 of them reported to be deceased. The SSN was available for 66.0% (311 of 471) of persons and in the case where the SSN was not available (30.4% of persons, 143 of 471) the date of death was known. In addition, 245 of the followed persons were LTFU and 87.3% (n=214) of them had a SSN. Overall, 361 of the 471 deceased patients and 7 of the LTFU patients could be linked to the SFSO death registry. Figure 1 summarizes the flow chart of the linkage between the SCCS and the SFSO. Among the linked persons in the SCCS, the reported date of death was identical for 298 persons, was different for 54 (for 11 the difference was ± 1 day), and was missing for 16 persons (including the 7 persons LTFU who could be linked).

**Fig. 1.**
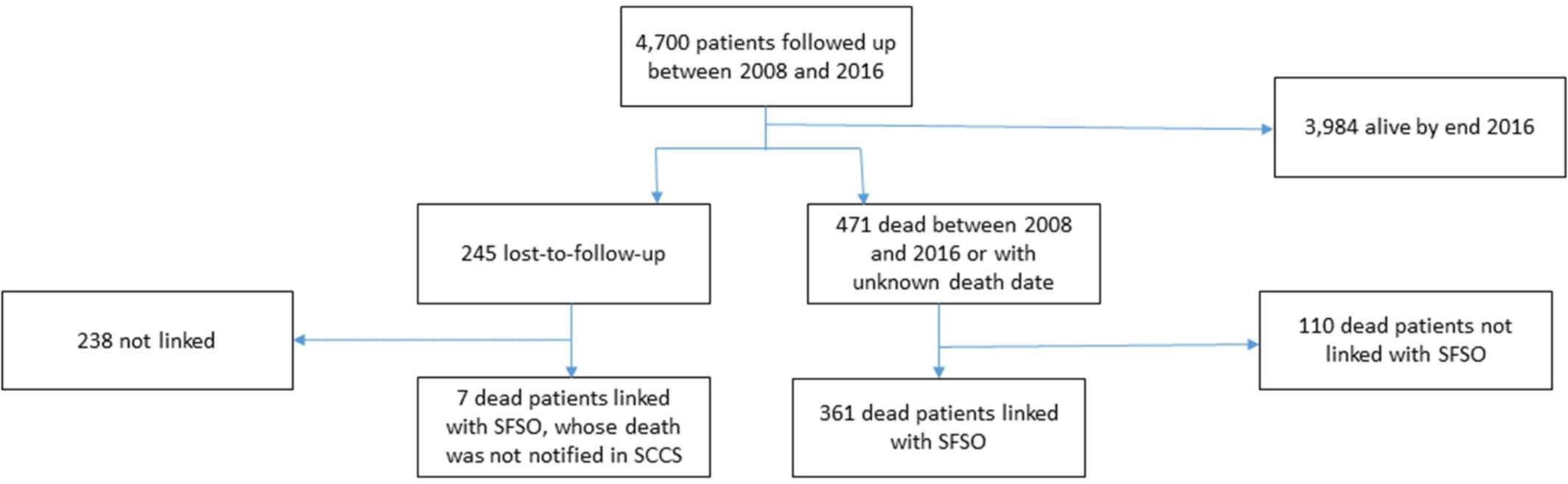
Flow chart illustrating the linkage between the Swiss Hepatitis C Cohort Study (SCCS) and the death registry of the Swiss Federal Office of Statistics (SFSO)

### Characteristics of persons in the SCCS

After linkage between the SCCS and the SFSO death registry, date of death was known for 474 persons and remained unknown for 4 persons. The median duration of follow-up per patient was 6.5 years (IQR: 2.2 - 9.0). During a total of 26,114 person-years (pyrs) of follow-up, the overall mortality rate was 18.3/1,000 pyrs (95%CI: 16.7 - 20.0).

Table 2 shows the baseline characteristics of all the SCCS persons who were included and of deceased persons who could or could not be linked to the death registry. Persons were mostly male (61.7%), Swiss nationals (73.5%), and their median age at start of follow-up was 47.5 years (IQR: 40.6 - 55.1). The majority (54.9%) had a history of IDU, 38.8% had received antiviral treatment, and 17.6% were cirrhotic. Deceased persons who could or could not be linked to the death registry had similar characteristics (Table 2).

**Table 2:**
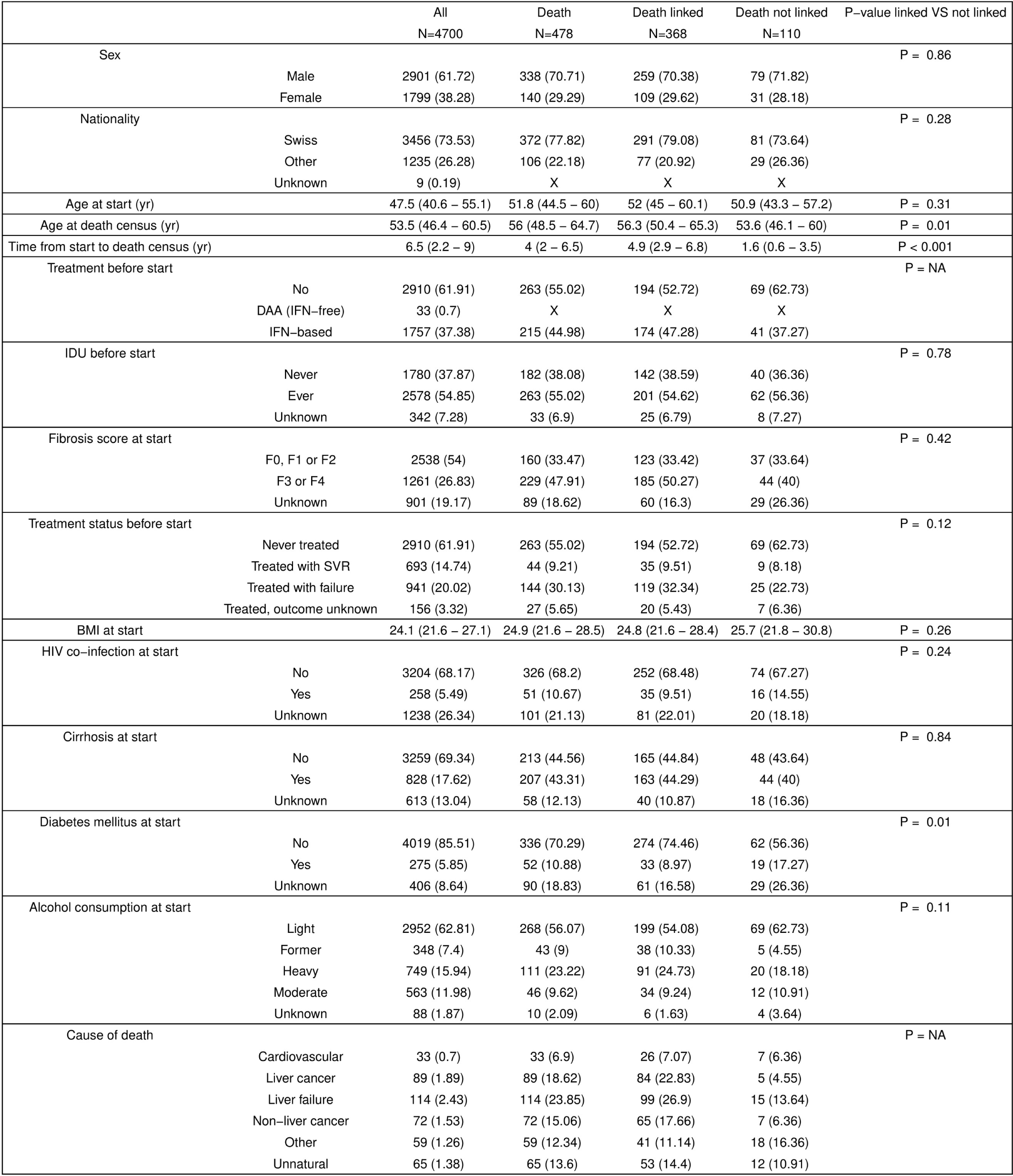
Characteristics of study participants at enrolment and causes of death in the Swiss Hepatitis C Cohort Study. P-values from Kruskal-Wallis rank sum test for continuous variables, and Chi-squared test for categorical variables are shown.

|  |  | All<br>N=4700 | Death<br>N=478 | Death linked<br>N=368 | Death not linked<br>N=110 | P-value linked VS not linked |
| --- | --- | --- | --- | --- | --- | --- |
| Sex |  |  |  |  |  | P = 0.86 |
|  | Male | 2901 (61.72) | 338 (70.71) | 259 (70.38) | 79 (71.82) |  |
|  | Female | 1799 (38.28) | 140 (29.29) | 109 (29.62) | 31 (28.18) |  |
| Nationality |  |  |  |  |  | P = 0.28 |
|  | Swiss | 3456 (73.53) | 372 (77.82) | 291 (79.08) | 81 (73.64) |  |
|  | Other | 1235 (26.28) | 106 (22.18) | 77 (20.92) | 29 (26.36) |  |
|  | Unknown | 9 (0.19) | X | X | X |  |
| Age at start (yr) |  | 47.5 (40.6 – 55.1) | 51.8 (44.5 – 60) | 52 (45 – 60.1) | 50.9 (43.3 – 57.2) | P = 0.31 |
| Age at death census (yr) |  | 53.5 (46.4 – 60.5) | 56 (48.5 – 64.7) | 56.3 (50.4 – 65.3) | 53.6 (46.1 – 60) | P = 0.01 |
| Time from start to death census (yr) |  | 6.5 (2.2 – 9) | 4 (2 – 6.5) | 4.9 (2.9 – 6.8) | 1.6 (0.6 – 3.5) | P < 0.001 |
| Treatment before start |  |  |  |  |  | P = NA |
|  | No | 2910 (61.91) | 263 (55.02) | 194 (52.72) | 69 (62.73) |  |
|  | DAA (IFN-free) | 33 (0.7) | X | X | X |  |
|  | IFN-based | 1757 (37.38) | 215 (44.98) | 174 (47.28) | 41 (37.27) |  |
| IDU before start |  |  |  |  |  | P = 0.78 |
|  | Never | 1780 (37.87) | 182 (38.08) | 142 (38.59) | 40 (36.36) |  |
|  | Ever | 2578 (54.85) | 263 (55.02) | 201 (54.62) | 62 (56.36) |  |
|  | Unknown | 342 (7.28) | 33 (6.9) | 25 (6.79) | 8 (7.27) |  |
| Fibrosis score at start |  |  |  |  |  | P = 0.42 |
|  | F0, F1 or F2 | 2538 (54) | 160 (33.47) | 123 (33.42) | 37 (33.64) |  |
|  | F3 or F4 | 1261 (26.83) | 229 (47.91) | 185 (50.27) | 44 (40) |  |
|  | Unknown | 901 (19.17) | 89 (18.62) | 60 (16.3) | 29 (26.36) |  |
| Treatment status before start |  |  |  |  |  | P = 0.12 |
|  | Never treated | 2910 (61.91) | 263 (55.02) | 194 (52.72) | 69 (62.73) |  |
|  | Treated with SVR | 693 (14.74) | 44 (9.21) | 35 (9.51) | 9 (8.18) |  |
|  | Treated with failure | 941 (20.02) | 144 (30.13) | 119 (32.34) | 25 (22.73) |  |
|  | Treated, outcome unknown | 156 (3.32) | 27 (5.65) | 20 (5.43) | 7 (6.36) |  |
| BMI at start |  | 24.1 (21.6 – 27.1) | 24.9 (21.6 – 28.5) | 24.8 (21.6 – 28.4) | 25.7 (21.8 – 30.8) | P = 0.26 |
| HIV co-infection at start |  |  |  |  |  | P = 0.24 |
|  | No | 3204 (68.17) | 326 (68.2) | 252 (68.48) | 74 (67.27) |  |
|  | Yes | 258 (5.49) | 51 (10.67) | 35 (9.51) | 16 (14.55) |  |
|  | Unknown | 1238 (26.34) | 101 (21.13) | 81 (22.01) | 20 (18.18) |  |
| Cirrhosis at start |  |  |  |  |  | P = 0.84 |
|  | No | 3259 (69.34) | 213 (44.56) | 165 (44.84) | 48 (43.64) |  |
|  | Yes | 828 (17.62) | 207 (43.31) | 163 (44.29) | 44 (40) |  |
|  | Unknown | 613 (13.04) | 58 (12.13) | 40 (10.87) | 18 (16.36) |  |
| Diabetes mellitus at start |  |  |  |  |  | P = 0.01 |
|  | No | 4019 (85.51) | 336 (70.29) | 274 (74.46) | 62 (56.36) |  |
|  | Yes | 275 (5.85) | 52 (10.88) | 33 (8.97) | 19 (17.27) |  |
|  | Unknown | 406 (8.64) | 90 (18.83) | 61 (16.58) | 29 (26.36) |  |
| Alcohol consumption at start |  |  |  |  |  | P = 0.11 |
|  | Light | 2952 (62.81) | 268 (56.07) | 199 (54.08) | 69 (62.73) |  |
|  | Former | 348 (7.4) | 43 (9) | 38 (10.33) | 5 (4.55) |  |
|  | Heavy | 749 (15.94) | 111 (23.22) | 91 (24.73) | 20 (18.18) |  |
|  | Moderate | 563 (11.98) | 46 (9.62) | 34 (9.24) | 12 (10.91) |  |
|  | Unknown | 88 (1.87) | 10 (2.09) | 6 (1.63) | 4 (3.64) |  |
| Cause of death |  |  |  |  |  | P = NA |
|  | Cardiovascular | 33 (0.7) | 33 (6.9) | 26 (7.07) | 7 (6.36) |  |
|  | Liver cancer | 89 (1.89) | 89 (18.62) | 84 (22.83) | 5 (4.55) |  |
|  | Liver failure | 114 (2.43) | 114 (23.85) | 99 (26.9) | 15 (13.64) |  |
|  | Non-liver cancer | 72 (1.53) | 72 (15.06) | 65 (17.66) | 7 (6.36) |  |
|  | Other | 59 (1.26) | 59 (12.34) | 41 (11.14) | 18 (16.36) |  |
|  | Unnatural | 65 (1.38) | 65 (13.6) | 53 (14.4) | 12 (10.91) |  |

### Comparison between causes of death in the SCCS and in the SFSO death registry

Of the 368 deceased persons who were linked to the death registry, the main cause of death was available from both the SCCS and the SFSO for 249 persons (67,7%), and for 227 (91,1%) of these 249 persons ICD-10 codes were provided in the SCCS. When comparing the causes of death using the ICD-10 codes, the Kappa statistic for interrater agreement was 0.14 (for 40 of 227 cases the codes were identical). If we used the grouped causes of death, the Kappa statistics increased to 0.45 (in 138 of 249 cases the causes were identical), demonstrating that the agreement was weak for both approaches.

### Time trends in all-cause and cause-specific mortality among the persons in the SCCS

Liver failure was the leading cause of death, with a crude death rate of 4.4/1000 pyrs (95% CI: 3.6 - 5.2), followed by liver cancer (3.4/1000 pyrs, CI 2.8 - 4.2), non-liver cancer (2.8/1000 pyrs, CI 2.2 - 3.4), unnatural causes (2.5/1000 p-years, CI 2.0 - 3.1), other causes (2.3/1000 pyrs, CI 1.8 - 2.9), and cardiovascular causes (1.3/1000 pyrs, CI 0.9 - 1.7). For 10% (n = 46) of the 478 deceased persons the main cause of death was unknown.

Figure 2 shows the evolution of crude mortality rates for all-cause mortality, liver failure and liver cancer mortality over the years. Similar curves for all the considered causes of death can be found in Appendix 2. Before 2014, the leading cause of death was liver failure, followed by liver cancer and non-liver cancer. In contrast, after 2014, when DAA became more widely available in Switzerland, liver cancer became the leading cause of death, followed by liver failure and non-liver cancer.

**Fig. 2.**
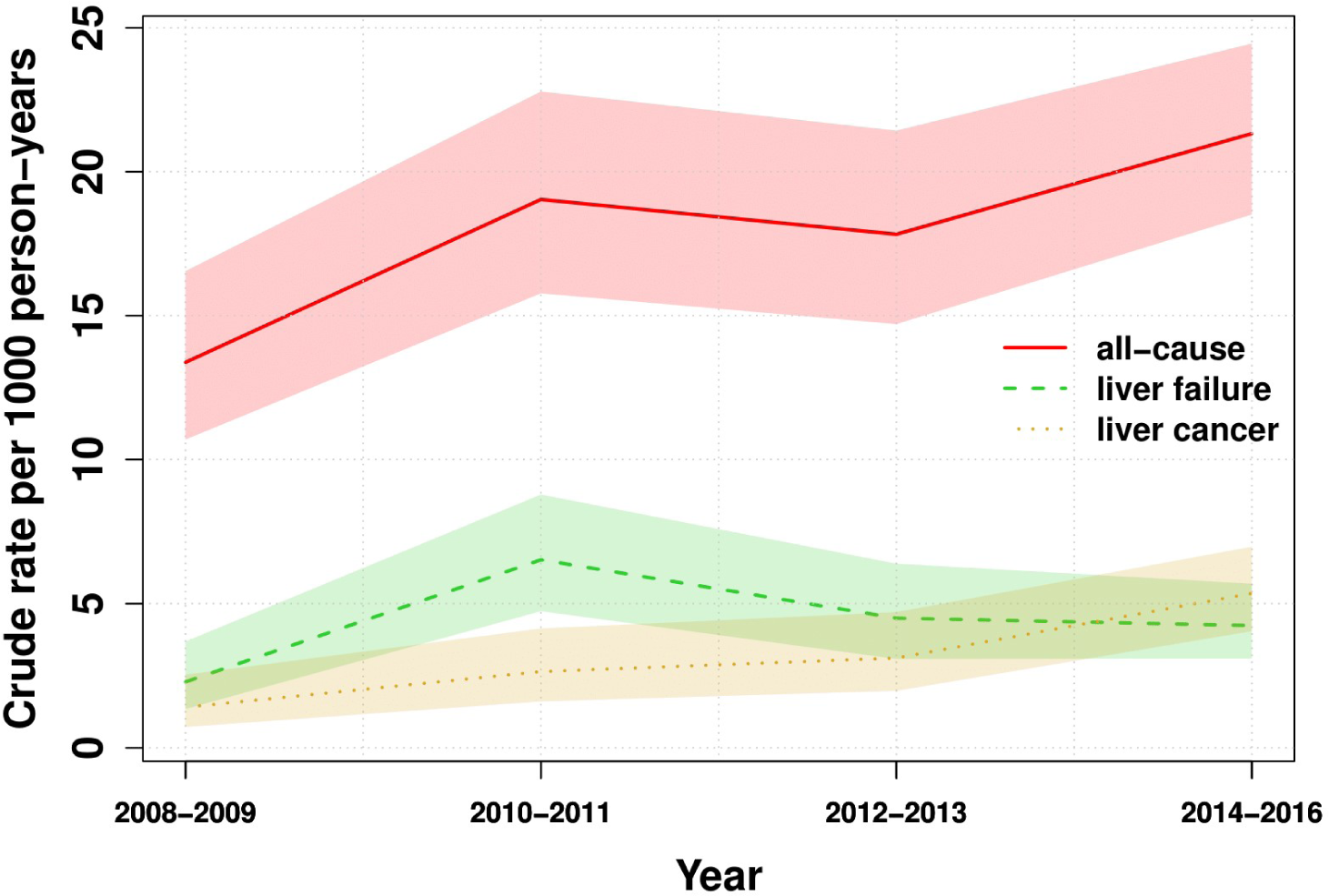
Crude mortality rates over the years for all-cause mortality, liver failure and liver cancer mortality in the Swiss Hepatitis C Cohort Study.

Figure 3 shows the cumulative incidence of causes of death over time. The overall probability of dying from any cause increased from 1.5% after one year of follow-up to 13.8% after eight years. While liver failure remained the most common cause of death throughout the follow-up period, liver cancer increased in relative proportion from year 6 onwards.

**Fig. 3.**
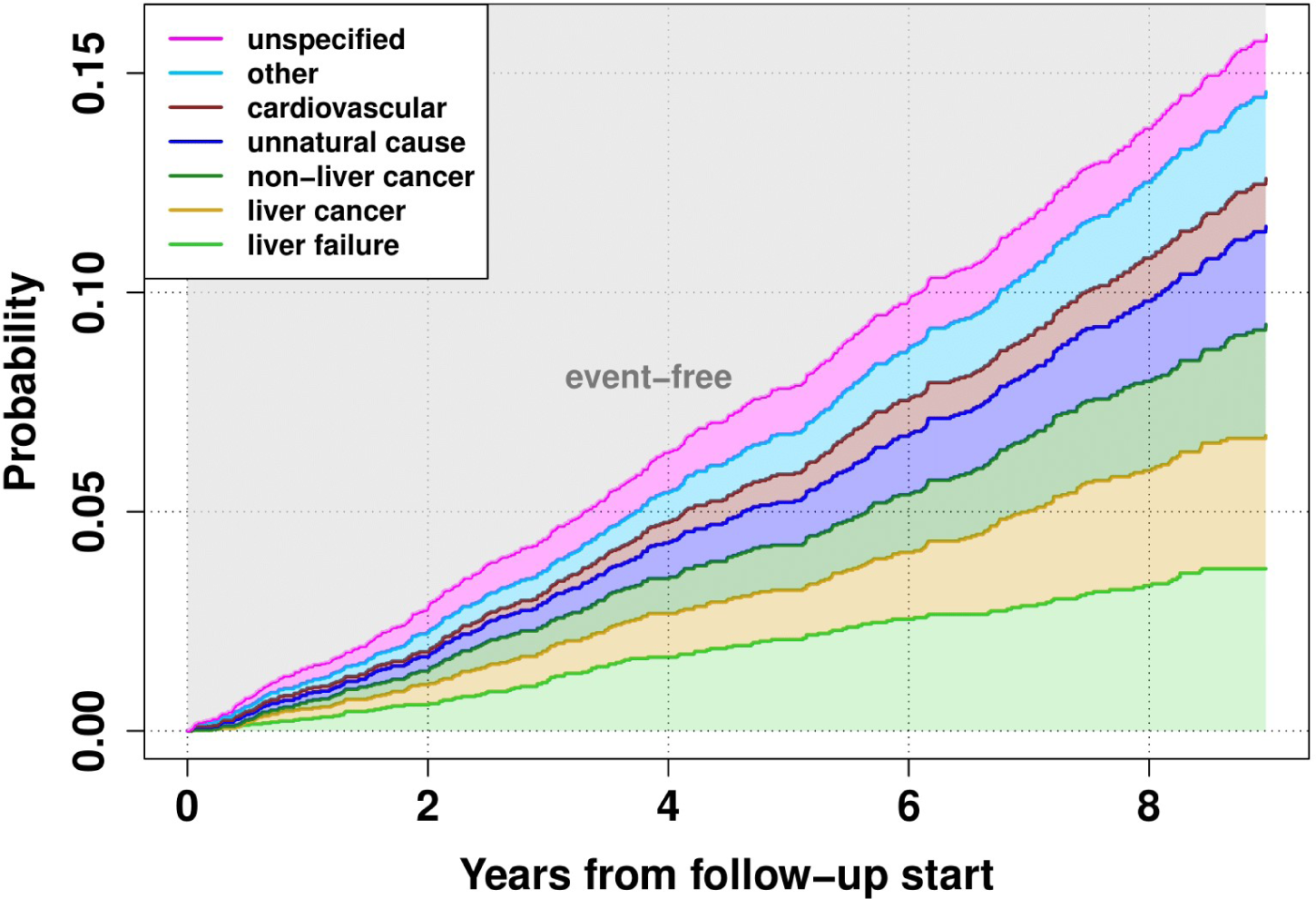
Cumulative incidence of different causes of death since registration into the Swiss Hepatitis C cohort study (causes of death are in the same order in the legend and on the plot)

Table 3 shows the risk factors for all-cause and cause-specific mortality in the SCCS over the period 2008-2016.

**Table 3:**
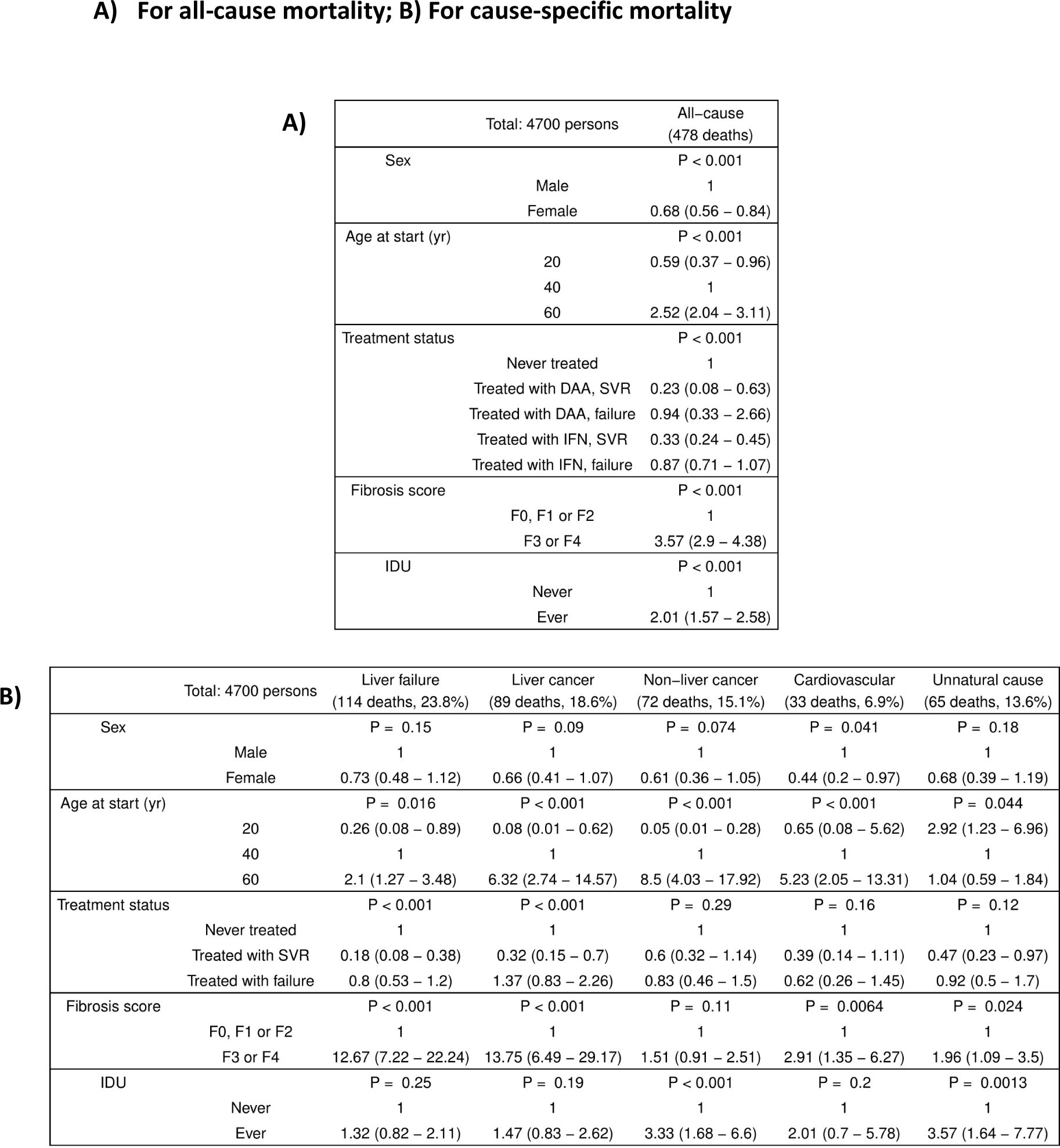
Estimates of the effect of gender, age, treatment, fibrosis stage and IDU on mortality, from Cox regression models, accounting for competing risk.

For all-cause mortality, the risk of death was increased for men, older persons, those with a high fibrosis stage and those with a history of IDU. Compared to persons who were never treated, the risk of death was lower for treated persons who reached SVR after receiving interferon (IFN)-based treatment (HR 0.33, 95% CI 0.24 - 0.45), and even slightly lower for treated persons who reached SVR after receiving DAA (HR 0.23, 95% CI 0.08 - 0.63). In contrast, treated persons who did not achieve SVR had a comparable risk of death to those who were never treated, irrespective of type of treatment.

Older persons and those with fibrosis score ≥ F3 were at higher risk of dying from liver failure than younger persons and those with fibrosis score ≤ F2. Baseline treatment status was associated with mortality due to liver failure, with a pronounced protective effect for treated persons with SVR, but no reduction in risk for persons treated without SVR.

For liver cancer, the risk of death was higher for older persons, as reported previously by other studies [20], and those with a high fibrosis stage. Having a history of IDU had little impact on liver cancer mortality. Compared to never treated persons, the risk of dying decreased for treated persons with SVR, but not for treated persons without SVR.

The risk of dying from non-liver cancer was higher for older persons than for younger persons. Although there was no significant association between non-liver cancer mortality and treatment status, nor with baseline fibrosis score, persons with a history of IDU were more at risk than persons without a history of IDU.

Having ever used injectable drugs was associated with an increased risk of dying from unnatural causes. In addition, persons aged 20 years were more at risk than persons aged 40 years or older, as well as persons with a fibrosis score ≥ F3 compared to those with fibrosis score ≤ F2. There was no association between unnatural causes of death and treatment status.

Results from the sensitivity analysis were similar to the main analysis for all-cause mortality, but there were some notable differences for cause-specific mortality (Appendix 3).

## Discussion

Based on the SCCS data, we investigated time trends and risk factors of all-cause and cause-specific mortality among HCV-infected persons in Switzerland. The linkage of the LTFU persons to the death registry did not significantly increase the number of deaths. However the proportion of unknown causes of death decreased substantially from 42% without linkage to 10% with linkage. The leading causes of death changed over time, with a substantial increase in the proportion of deaths related to liver cancer. Two factors probably explain this change: the risk of dying from liver disease increased as people became older; and ascertainment of cause of death may have improved over time. Indeed, the proportion of unspecified causes of death decreased from 13% in 2008-2013 to 4% in 2014-2016, reflecting the better linkage in the latter years (92% of deceased persons could be linked in 2014-2016, compared to 67% in 2008-2013).

Similarly to other studies [7], we found that mortality remained stable among HCV-infected persons in Switzerland over the past few years. The expected decrease in mortality due to DAA is therefore not occurring, or more probably not yet observable. The risk of death was lower for women, younger persons, persons with lower fibrosis score, persons without a history of IDU, and treated persons who reached SVR, with a first indication of a more positive effect with DAA compared to IFN-based treatments. However, in contrast to some other studies, we found little evidence that being treated with subsequent treatment failure had a protective effect compared to remaining untreated [19, 20].

For both liver failure and liver cancer, the risk of dying was significantly associated with treatment, with a lower risk for treated persons with SVR than for treated persons without SVR or for untreated persons. As data for wider use of DAA was only available for three years (2014 - 2016), we were not able to evaluate the difference between successful IFN treatment and successful DAA treatment with respect to liver-related mortality. Furthermore, in the early DAA era, the reimbursement of DAA by health insurances was limited to persons with advanced fibrosis (Metavir stages F3 or F4). By mid-2016, patients with Metavir score F2 were also treated [21, 22]. The limitation based on fibrosis stage was relaxed progressively between July and October 2017, when all patients were treated, although the prescription of DAA remained restricted to specialists.

Having a history of IDU was an important risk factor for death from non-liver cancer and unnatural causes of death, explained by the inclusion of overdose/accidental poisoning in this category. Persons who inject drugs (PWID) are a key subgroup of HCV-infected persons in Switzerland and other high-income countries, where the majority of HCV infections occurred via injection of drugs [23, 24].

Despite the implementation of the four-pillar strategy [27], a national drug policy in Switzerland since 1994, additional obstacles exist for HCV treatment among PWID both from treatment providers and from patients [27]. This results in some HCV-infected PWID entered care relatively late, at an advanced fibrosis stage. The proportion of infected PWID who received anti-HCV treatments increased only slightly from 51% in the years 2008-2013 to 58% in the years 2014-2016. The delay in treatment initiation probably decreased, and even PWID who enter care late are more likely to reach favourable outcomes with DAA. However, persons with a history of IDU now have a higher risk of dying from non-liver cancer, potentially related to their addiction and to additional risk factors that are common among PWID, such as smoking or excessive alcohol consumption.

Differences between causes of death reported in the SFSO death registry and the SCCS can be partly explained by different coding practices. Detailed analysis of the discrepant entries revealed that for 19% of cases where the cause of death was coded as “other” in the SCCS, it was coded as “non-liver cancer”, “unnatural cause”, or “cardiovascular disease” in the SFSO. The main focus of the SCCS was likely to identify all HCV-related deaths, and there was less emphasis to record the exact cause of death. For 21% of SCCS entries, cause of death was “liver failure” in one dataset and “liver cancer” in the other, two causes that are relatively similar. Overall, clear discrepancies in the causes of death recorded in the SCCS and the SFSO existed in only 21% of cases (Appendix 4).

To our knowledge, this is one of the first studies investigating detailed risk factors and time trends of cause-specific mortality among HCV-infected persons. Due to the linkage with the death registry, we significantly improved information on causes of death. By accounting for competing risks, by including age as a continuous covariate (and not categorizing it arbitrarily), and by adjusting models for time-dependent covariables, we could improve the accuracy of our analysis.

A limitation of this study was the unavailability of complete data relating to fibrosis stage evolution. Due to this, we made no assumption on the evolution of fibrosis over time, and used multiple imputation to impute missing values, which may compromise the robustness of our analysis. We did a sensitivity analysis, using complete cases only. In this analysis, the number of persons for the cause-specific analysis was relatively limited and results may therefore be less reliable.

In conclusion, although mortality did not, or did not yet decrease with DAA, we found that causes of death changed over time. With the wider use of DAA, irrespective of fibrosis stage, it is likely that liver-related mortality will continue to decline in the future. Continuous monitoring of mortality and causes of death will therefore remain important to evaluate the long-term effect of DAA and to develop effective interventions.

## Data Availability

The data referred to in that manuscript is not publicly available.

## Data Availability

The data referred to in that manuscript is not publicly available.

## Data Availability

The data referred to in that manuscript is not publicly available.

## Acknowledgments

The Swiss Hepatitis C Cohort Study group comprises Francesco Negro, Laurent Kaiser (Geneva); Markus Heim, Hans Hirsch (Basel); Nasser Semmo, Franziska Suter (Bern); Darius Moradpour, Vincent Aubert (Lausanne); Hans H. Siegrist (La Chaux-de-Fonds); Andreas Cerny, Gladys Martinetti Lucchini (Lugano); Olivier Clerc (Neuchâtel); David Semela, Patrick Schmid, Günter Dollenmaier (St. Gallen); Beat Müllhaupt, Elsbeth Probst-Müller (Zürich); Pascal Benkert, Thomas Fabbro, Marielle Rutquist, Constantin Sluka (Basel Clinical Trial Unit).

We gratefully thank the SCCS study nurses: Ulrike Schnee-Lach (Basel); Kathrin Husi, Alice Gilg (Bern); Nicole Eicher, Fabio Cassano (Geneva); Adeline Mathieu, Maribelle Herranz (Lausanne); Claudia Dibartolomeo (Lugano); Marielle Grosjean (Neuchâtel); Patrizia Künzler-Heule, Roman Stillhard, Michelle Grahornig, Simone Kessler (St. Gallen); Barbara Brunner-Geissmann, Andrea Zumbühl (Zürich).

We thank the Swiss Federal Statistical Office for providing mortality data and for their support, which made this study possible.

We thank Danny Sheath and Rachel Esra for the editorial support.

## Appendix

### Appendix 1: Details on the multiple imputation

Because adding variables that are not part of the analysis can improve the imputation, we added the following baseline explanatory variables: body mass index (BMI), HIV co-infection, cirrhotic status, diabetes comorbidity, alcohol consumption (light/no drinker if average consumption ≤ 20g/day, moderate drinker if average consumption between 21 and 40g/day, excessive drinker if average consumption > 40g/day, and former drinker if ever drank more than 40g/day but is now a light or moderate drinker). Furthermore, the outcome was also included in the imputation. We ran the model on 20 imputed datasets for each analysis and combined the estimates with Rubin’s rule.

### Appendix 2: Time trends in mortality rates (with 95% confidence intervals), per cause of death

The x-axis corresponds to the year of death

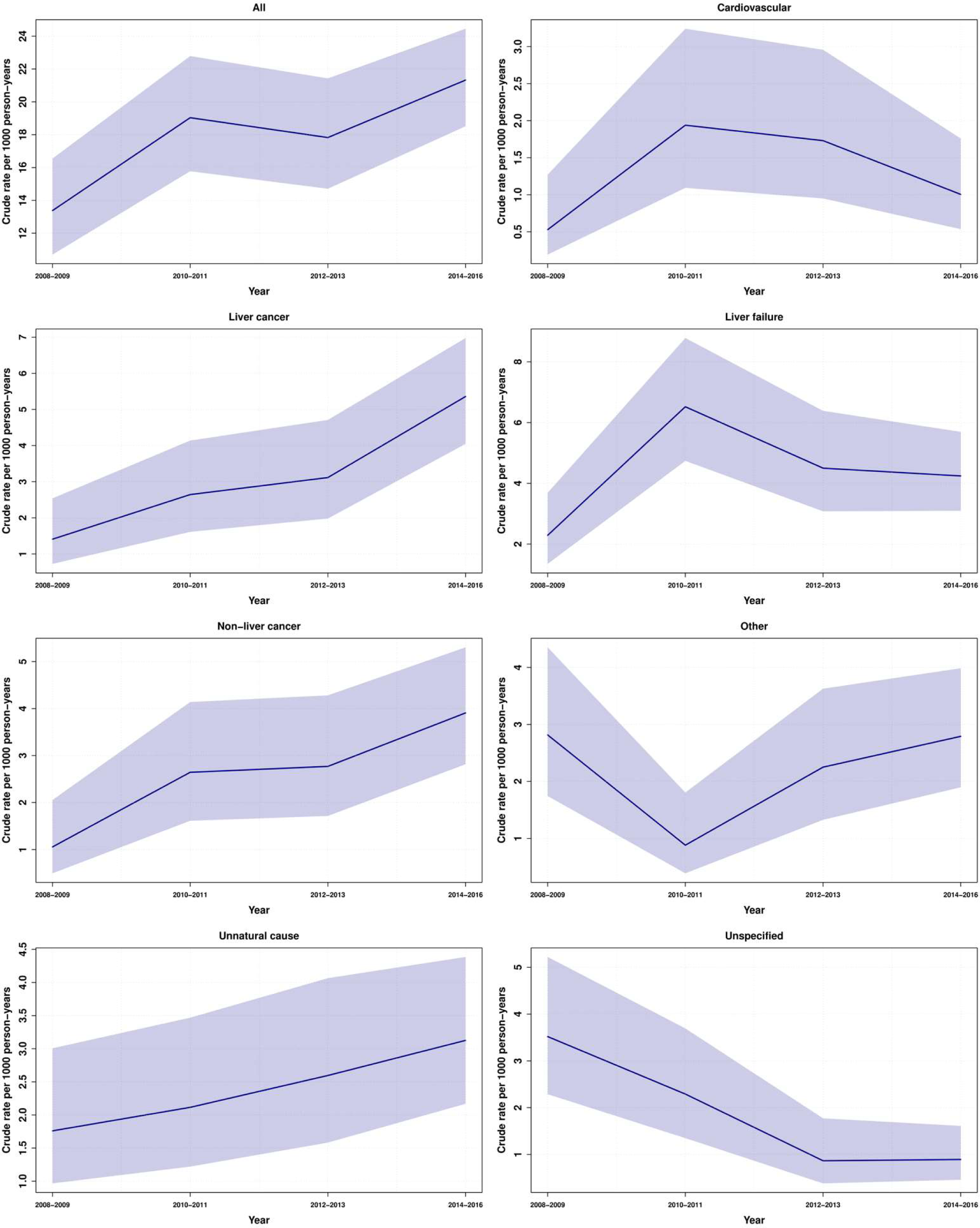

### Appendix 3: Hazard ratios from Cox regression models, accounting for competing risk, excluding persons with missing information (sensitivity analysis)

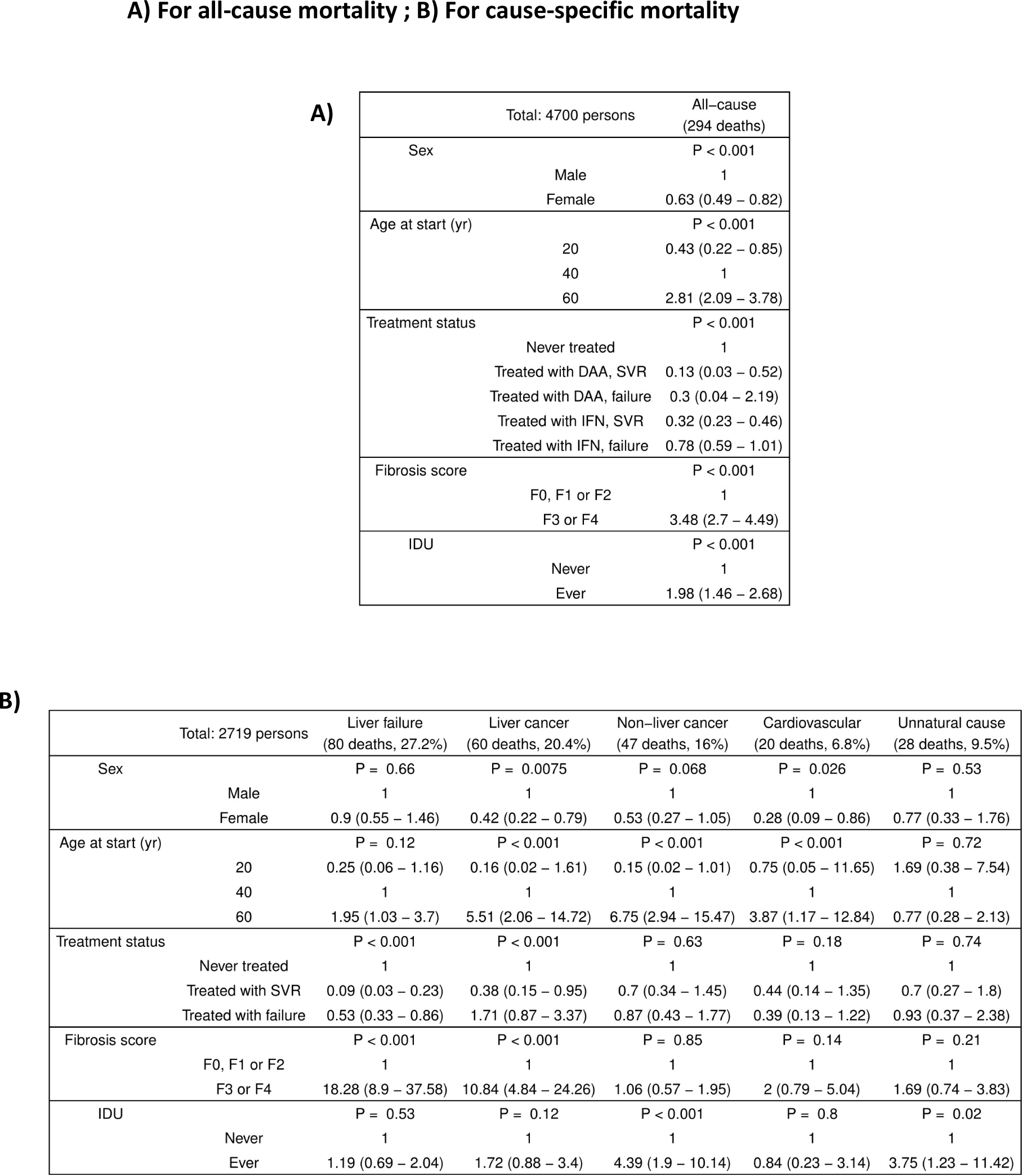

### Appendix 4: Comparison between SCCS and SFSO death cause groups

| SCCS / SFSO | Cardiovascular | Liver cancer | Liver failure | Non–liver cancer | Other | Unnatural cause |
| --- | --- | --- | --- | --- | --- | --- |
| Cardiovascular | 9 | 2 | 9 | 3 | 3 | 1 |
| Liver cancer | 0 | 34 | 1 | 2 | 0 | 0 |
| Liver failure | 3 | 22 | 34 | 2 | 7 | 1 |
| Non–liver cancer | 0 | 2 | 1 | 33 | 0 | 0 |
| Other | 6 | 6 | 21 | 12 | 14 | 3 |
| Overdose | 1 | 0 | 0 | 0 | 1 | 5 |
| Unnatural cause | 0 | 0 | 0 | 0 | 2 | 9 |
| Unspecified | 10 | 13 | 24 | 15 | 29 | 28 |

